# Repurposing Therapeutics for COVID-19: Rapid Prediction of Commercially available drugs through Machine Learning and Docking

**DOI:** 10.1101/2020.04.05.20054254

**Authors:** Sovesh Mohapatra, Prathul Nath, Manisha Chatterjee, Neeladrisingha Das, Deepjyoti Kalita, Partha Roy, Soumitra Satapathi

## Abstract

**Background:** The outbreak of the novel coronavirus disease COVID-19, caused by the SARS-CoV-2 virus has spread rapidly around the globe during the past 3 months. As the virus infected cases and mortality rate of this disease is increasing exponentially, scientists and researchers all over the world are relentlessly working to understand this new virus along with possible treatment regimens by discovering active therapeutic agents and vaccines. So, there is an urgent requirement of new and effective medications that can treat the disease caused by SARS-CoV-2.

**Methods and findings:** We perform the study of drugs that are already available in the market and being used for other diseases to accelerate clinical recovery, in other words repurposing of existing drugs. The vast complexity in drug design and protocols regarding clinical trials often prohibit developing various new drug combinations for this epidemic disease in a limited time. Recently, remarkable improvements in computational power coupled with advancements in Machine Learning (ML) technology have been utilized to revolutionize the drug development process. Consequently, a detailed study using ML for the repurposing of therapeutic agents is urgently required. Here, we report the ML model based on the Naïve Bayes algorithm, which has an accuracy of around 73% to predict the drugs that could be used for the treatment of COVID-19. Our study predicts around ten FDA approved commercial drugs that can be used for repurposing. Among all, we suggest that the antiretroviral drug Atazanavir (DrugBank ID – DB01072) would probably be one of the most effective drugs based on the selected criterions.

**Conclusions:** Our study can help clinical scientists in being more selective in identifying and testing the therapeutic agents for COVID-19 treatment. The ML based approach for drug discovery as reported here can be a futuristic smart drug designing strategy for community applications.

**Author summary:** *Why was this study done?:*

- The recent outbreak of novel coronavirus disease (COVID-19) is now considered to be a pandemic threat to the global population. The new coronavirus, 2019-nCoV has now affected more than 200 countries with over 17,83,941 cases confirmed and 1,09,312 deaths reported all over the world [as on 12 April 2020].
- There is an urgent need for the development of drugs or vaccine which can save people worldwide. However, the vast complexity in drug design and protocols regarding clinical trials often prohibit developing various new drug combinations for this epidemic disease. Recently, Artificial Intelligence (AI) technology have been utilized to revolutionize the drug development process. Can we use AI based repurposing of existing drugs for accelerated clinical trial in the treatment of COVID-19?

*What did the researchers do and find?:*

- Here, we report the Machine Learning (ML) model based on the Naïve Bayes algorithm, which has an accuracy of around 73% to predict the drugs that could be used for the treatment of COVID-19.
- Our study predicts around ten FDA approved commercial drugs that can be used for repurposing. Among all, we suggest that the antiretroviral drug Atazanavir (DrugBank ID – DB01072) would probably be one of the most effective drugs based on the selected criterions.

*What do these findings mean?:*

- The present approach will save a lot of resources and time for synthesizing novel drugs and thus will be useful for a vast majority of medical research community.

## Introduction

The recent outbreak of novel coronavirus disease (COVID-19) is now considered to be a pandemic threat to the global population[1–3]. Coronaviruses belong to a family of viruses mainly found in animals but with the recent outbreak, they have transmitted to humans. The new coronavirus, 2019-nCoV is termed as severe acute respiratory syndrome-related coronavirus SARS-CoV-2[4–9] which has now affected more than 200 countries with over 17,83,941 cases confirmed and 1,09,312 deaths reported all over the world [as on 12 April 2020]. This could potentially bring major challenges to global healthcare and disastrous effect on the global economy if the virus is not contained within a few months[10]. The common symptoms include cough, fever, shortness of breath, fatigue etc which makes it confusing for the patients to differentiate the symptoms with that of the typical cold and flu[10–13]. Reports suggest that the virus is transmitted through body fluids of the infected patients, especially when in contact and while sneezing even though exact reasons are not known. Unfortunately, no drugs have been approved by regulatory agencies to treat SARS-CoV-2 infection until now. Efforts are ongoing on war footing to find the effective drug and vaccine to treat this pandemic.

Coronaviruses are classified into four classes designated as alpha, beta, gamma, and delta[14]. The betacoronavirus class includes severe acute respiratory syndrome virus (SARS-CoV), Middle East respiratory syndrome virus (MERS-CoV), and the COVID-19 virus (SARS-CoV-2). Coronaviruses are found to be considerably large viruses with a single-stranded positive-sense RNA genome encapsulated inside a membrane envelope having proteins apprearing like spikes protruding from their surface. These spikes adheres onto human cells, through certain receptors on target cells, after which undergoes a structural change that lets the viral membrane fuse with the cell membrane. The viral genes then enter the host cell, producing more viruses. Recent studies show that, like the virus responsible for 2002 SARS outbreak, SARS-CoV-2 spikes also bind to receptors on the human cell surface called angiotensin-converting enzyme 2 (ACE2)[15]. Like SARS-CoV and MERS-CoV, SARS-CoV-2 also attacks the lower respiratory system causing viral pneumonia. However, there are also reports that it could affect the gastrointestinal system, heart, kidney, liver, and central nervous system resulting in multiple organ failure[16]. Compiling the medical reports and data available from the patients, SARSCoV-2 is found to be more transmissible/contagious than SARS-CoV[17].

Rapid development of computer aided technology like ML based on Artificial Intelligence (AI) can help accelerate the drug development process for different diseases[18–20]. The advantage of AI approaches like ML is that they can be applied to learn from examples and build predictive models even when our understanding of the underlying biological processes is limited, or when computational simulations based on fundamental physical models are too expensive to be carried away. Another advantage of ML is to automatically learn to identify complex patterns that categorize sets from input data and thereby make intelligent decisions based on independent datasets[21]. ML can accurately predict drug-target interactions as an enormous amount of complex information by studying hydrophobic interactions, ionic interactions, hydrogen bonding, van der Waals forces, etc. between molecules. Bioactivity datasets which are available from the numerous high throughput screens deliver useful means for machine learning classifiers as they contain binary information (active/inactive) as well as numerical values to classify different compounds under consideration[22,23]. Such a huge number of datasets available on biological activities of molecules, derived from high throughput screens now allows to create predictive computational models.

In this study, we have applied a machine learning approach to predict several new potential drugs for the treatment of SARS-CoV-2 and validated the predicted drugs. Initially, we have trained our model with the inhibitors of the *SARS Coronavirus 3C-like Protease*. The FDA approved drugs are only taken from the Drug bank as a test model to predict the new drugs. These new drugs are again validated using a docking method to ensure that the drugs match with the same active site on the protein. A ranked list of drugs based on energy value is given that can be tested experimentally. Our study hypothesizes that the commercial FDA approved antiretroviral drug Atazanavir may be a potential candidate requiring to limit viral recognition of host cells or disrupt host-virus interactions thus requiring further clinical trial.

## Methods

### Preparation of Dataset

In the present study, the compounds of the dataset are tested in the cell based system using plate readers and then their results are stored as Bioassay Dataset in the Pub chem. This dataset of PubChem Bioassay assigned AID 1706 contains around 290893 compounds as one activity set and they are the inhibitors of *SARS coronavirus 3C-like Protease* in the cells. This dataset is stored in the section Bioassay of PubChem database of National Centre for Biotechnology Information (NCBI), and they have the identification AID number as AID 1706[23]. This corresponding bioassay belongs to the Scripps Research Institute Molecular Screening Center of replication in *SARS coronavirus 3C-like Protease* in the cells. The compounds are classified under three distinct categories as actives, inactives and inconclusive. Compounds that inhibit luminescence activity may kill *SARS coronavirus*, inhibit *SARS coronavirus* invasion or inhibit development of the parasite within the host cell and hence these are classified under the active section and the compounds which do not show effectiveness are classified under inactive section. These complete datasets were downloaded in the form of SDF (Structure Data File) from the PubChem Database.

The Drug Bank is an online database which contains detailed data about various medications[25]. Today, it is being widely used to facilitate in silico drug target discovery, drug design, drug docking or screening, drug metabolism prediction, drug interaction prediction and for general pharmaceutical education. This database of more than 4900 Drugs is categorized into many different types as Trial stages Drugs, Approved Drugs and Withdrawn Drugs. In this database, more than 45% of drugs are approved for various medication purposes[25]. In this research, we have focused only on the FDA approved drugs for repurposing purpose which are around 2388 with the intention that it will minimize clinical trial in the present situation. These drugs were downloaded in the form of SDFs and after processing, the descriptions generated were taken as the test model for developing the train model which was made on the basis of a database containing the inhibitors of the *SARS coronavirus*. The developed model has predicted few of the potential drugs. The NCBI Protein Database was used in the process of getting the FASTA sequence of the desired protein (*SARS coronavirus*). Further, the FASTA sequence is used for the modeling of the three-dimensional protein structure and on the basis of this structure the docking of the known and predicted drugs have been carried out[26].

### Processing Dataset

Since the datasets are present in the form of SDFs, we have generated the attributes present in the SDFs. First, the information present in the SDFs are generated as CSV files which are used as the training dataset and test dataset for preparing the ML models. These CSV files containing both the actives and inactive points are split into 80% as training dataset and 20% as test dataset. This entire splitting process was random. This process is done by self-written python code to split as per the conditions.

### Classification Algorithm

We have used Machine learning (ML) model to the selected dataset from the PubChem which was considered as inhibitors and tested against the drugs from the Drug Bank to find more suitable drugs for the CoronaVirus-19[27]. Using ML, we have implemented the classification algorithms as described below.

The classification is a type of supervised learning in which the computer system can learn from the dataset which contains the detail and practical results. The algorithmic procedure of the classification is to assign an input value according to the description in the datasets[28]. So, for this, it requires a mathematical classifier that can assign specific class (active and inactive) labels to instances defined by the attributes. In this process, the training model is made to learn using dataset where the classification is already assigned and on the basis of which it is able to run on different datasets to classify them according to the present instances. In this study, we have compared the results from the classifier that is Naïve Bayes classification algorithm.

Naive Bayesian classification algorithm is a simple and elegant approach by assuming that its classification attributes are independent and they don’t have any correlation with each other[29]. It is a type of classifier that depends on Bayes’ hypothesis. Naive Bayes does work best in two cases: complete independent feature (as expected) and functionally dependent features (as expected) and is a widely tested method for probabilistic induction. This classifier functions well and has advantages over many other induction algorithms. It has no entangled iterative parameter that makes it work for vast data sets.

This algorithm is more useful than any other induction algorithms because of its computation speed and reliability. It can be useful for both the binary classification as well as multi-classification ^29^.

### Training Model

The training model is prepared by 80% of the original dataset. The dataset is completely classified from where the computer learns and finds the relations among various attributes. The cross-validation is used along with the algorithm to train the model. In this case, the cross-validation is n set with n-folds dataset. Then it is supposed to divide the training dataset into n parts, and the n-1 parts will be used as training data and the other one will be used to validate the rest. This process of iteration goes on for n iteration times. Here, we have used 10-fold and it is chosen as per the size of the dataset[30,31].

Generally, the datasets containing binary classification based on several attributes are imbalanced. We observe the similar tred here. These imbalanced datasets are not possible to be handled by the normal classifiers since they give importance to each of the attributes equally which could lead to misclassification errors. This can decrease the accuracy of the dataset for the trained model[32,33]. Therefore, we have used the misclassification cost where the trained model becomes cost sensitive and able to find the lowest expected cost. This approach is actually much randomized because it neither depends upon the number of attributes nor on the minority class ration; rather it depends on the base classifier[34,35].

Here, we had two methods to introduce the misclassification cost with the imbalance dataset. The first method is to classify the algorithm into the cost-sensitive one and proceed with the rest settings[36]. The other is the use of a wrapper, which helps in the base classifiers into cost sensitive ones.

We have used Naïve Bayes classifier which uses the cost insensitive algorithm to predict the probability estimations of the test instances and then using this it predicts class labels for the examples of the test dataset. In our report, we have classified our datasets into two classes i.e. active and inactive. So, we used the 2X2 matrix which is generally used for the binary classification. In the matrix sections are True Positives (active classified as active), False Positives (Inactive classified as active), False Negatives (active classified as inactive) and True Negatives (inactive classified as inactive). In this case, the percent of False Negatives are more important than the percent of False Positives and the upper limit for False Positives were set to 20%[32,36]. In this process, we increase the misclassification up to the set percent which also helps in the increasing of the True Positives.

Since, the actives are very less in number, we have replicated them to around 100-110 times to match it with the inactives and make the model less biased.

### Independent Validation

There are various methods for the validation of the binary classifiers. The True Positive Rate is the ratio of the actual actives to the predicted positives and this can be obtained as (TP/TP+FN). The False Positive Rate is the ratio of the predicted false actives to actual inactives and this can be obtained as (FP/TN+FP). Accuracy shows the model’s performance relative to the real values and this can be calculated as (TN+TP/TN+TP+FP+FN). The Sensitivity shows the model’s ability to identify the positive results and this is calculated as (TP/FN+TP) and the Specificity shows the model’s ability to identify the negative results and this is calculated as (TN/TN+FP). A model with high specificity and sensitivity has a low error rate. The Balanced Classification Rate (BCR) is the mean of the sensitivity and specificity, which provides the accuracy of the model applied on the imbalanced dataset. This BCR can be calculated as 0.5*(specificity+sensitivity).

Apart from the BCR, the Mathews Correlation Coefficient (MCC) is also used whose range varies from -1 to 1. The Receiver Operating Characteristic (ROC) curve is the visualization of the ratio of FPR to TPR. In this case, the FPR and TPR are placed on the x- and y-axis respectivel. The Area under curve shows the probability prediction of the classifier and its ability to classify the randomly chosen instance into the correct class.

### Docking of the Predicted Drugs

Around 178 drugs were predicted by our ML model which can be effective for the treatment of diseases caused by *SARS-Cov-2*. There are no available drugs as of now, since the epidemic has just recently accelerated to over 12,18,991 cases [As of 5th April, 2020].

The predicted compounds with above 95% of confidence were docked using patchdock web server.

## Results

Here, we have at first taken the inhibitors of SARS-CoV-2, which doesn’t allow them to replicate in the host. These are screened and collected in the bioassay AID 1706 which were used as the main component for the modeling of the training model using ML. The 914 attributes were taken under consideration for more than 200,000 compounds. We have not used unsupervised learning to filter out the dataset, because it would have made the dataset much weaker. As mentioned in the method section, we have used a classifying algorithm to train the model and the best among them was further used for the testing and predicting the drugs from the Drug Bank. The schematic of the process is shown in Figure 1.

**Figure 1:**
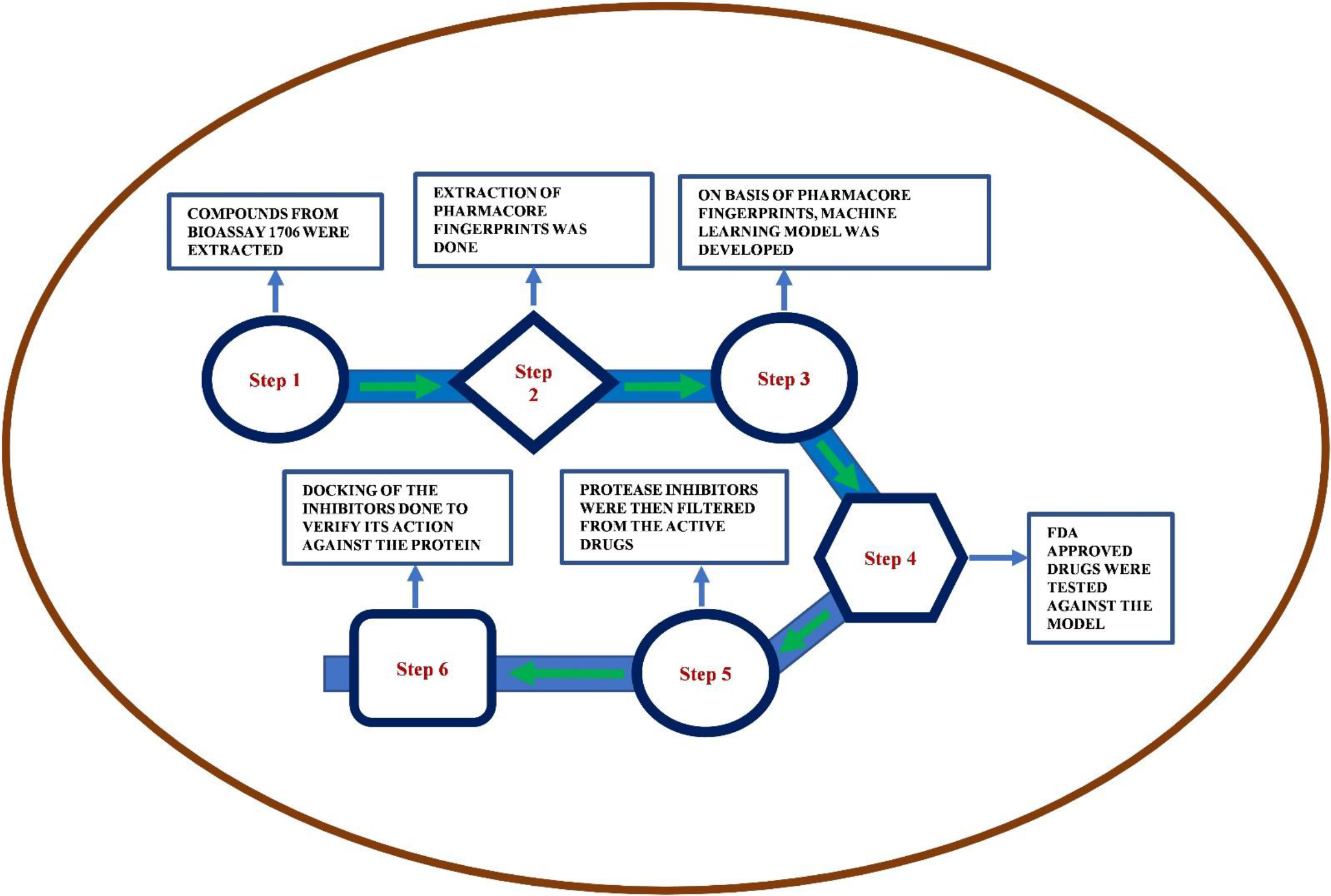
Flowchart of the process.

We have used the Naïve Bayes against the dataset. It has shown accuracy of approximately 72.999%. With this model, we have used the drugs from the Drug Bank to get predicted for the identification of the potential drugs which can be used for the treatment of disease caused by *SARS coronavirus*. Along with that the model has predicted 34754 True Positives and 3904 True Negatives.

The Naive Bayes has 0.194 MCC and BCR i.e., 76.69%. As per the independent validation, the algorithm with the lowest possible False Positives and highest possible True Positives can be considered to be the most effective model for the prediction of the drugs from the Drug Bank. In all the cases, the compounds for the False Positives were set to 23%. The comparison of the False Positive Rate and True Positive Rate for all algorithms used in the case of preparation of ML models are shown in Figure 2. When these results are compared with the rest of the dataset, it is found to be way better because it has satisfied both criteria and the rest of the algorithms has not reached the mark that has been achieved by Naive Bayes.

**Figure 2:**
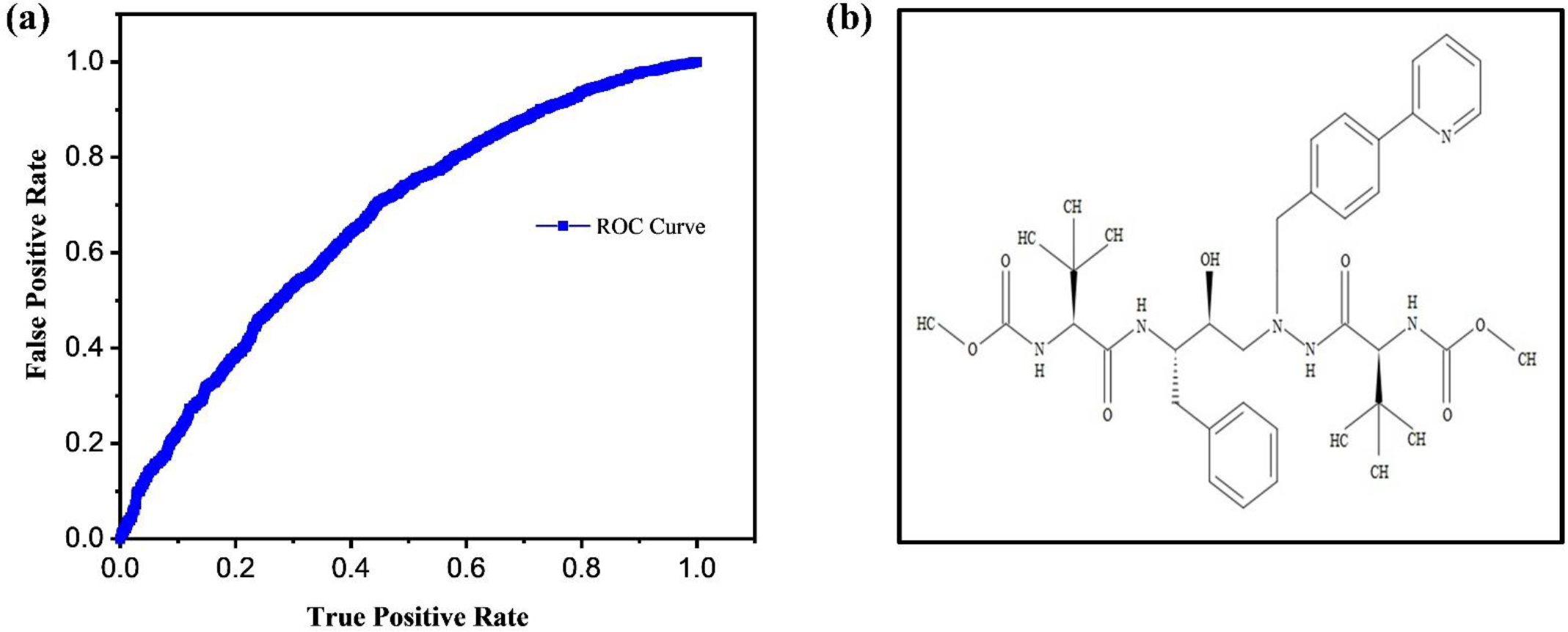
(a) The comparison of the False Positive Rate and True Positive Rate for all Algorithms used in case of preparation of Machine Learning models (b) Chemical Structure of the drug.

The model created with Naive Bayes algorithm predicted around 471 drugs out of all the 2388 approved drugs. These 471 drugs contain all the drugs which may be for the treatment of disease caused by *SARS coronavirus-19*. Here, we have predicted several drugs and out of which, we have docked and suggested the top 10 of them keeping align both the ML accuracy and the docking result. The docking results have been used for binding energy prediction and its effectiveness to bind with the compound. The results are shown in Table 1.

**Table 1:**
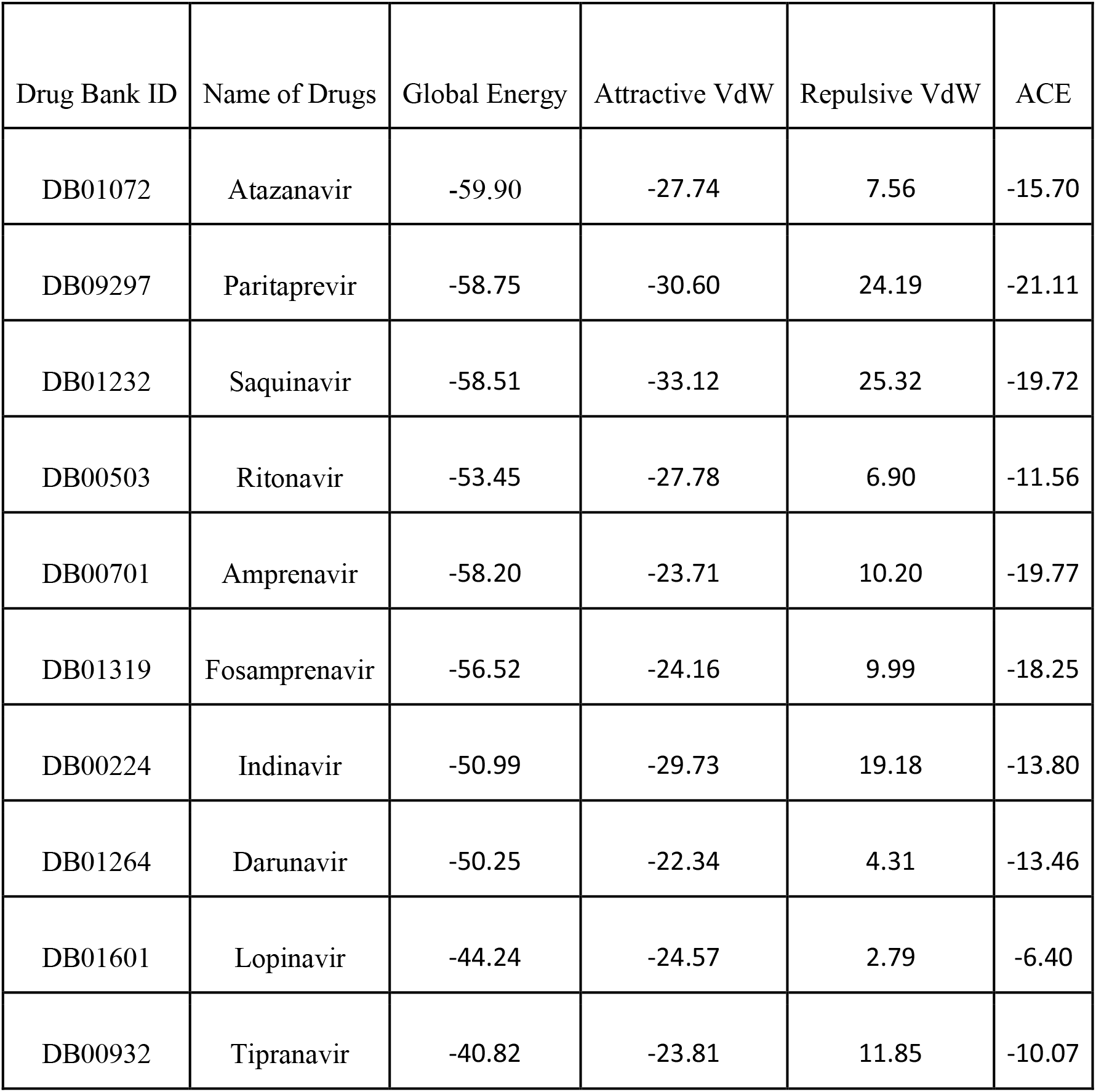
Global energy value for predicted drugs

Based on the docking study, we identified 10 possible drugs namely Atazanavir (DrugBank ID – DB01072), Paritaprevir (Drug Bank ID: DB09297), Saquinavir (Drug Bank ID: DB01232), Ritonavir (Drug Bank ID: DB00503),, Amprenavir (Drug Bank ID: DB00701), Fosamprenavir (Drug Bank ID: DB01319), Indinavir (Drug Bank ID: DB00224), Darunavir (Drug Bank ID: DB01264), Lopinavir (Drug Bank ID: DB01601) and Tipranavir (Drug Bank ID: DB00932) for the treatment of novel SARS Coronavirus.

The PDB structure of SARS-CoV 3C-like protease was retrieved from Protein data bank (PDB ID: 3VB7). Intitially, the strcute was optimized by removing water moplecules and other ligand. All the ligands structrure were retrived from pubchem (https://pubchem.ncbi.nlm.nih.gov/). The docking experiment was done with the patchdock server (https://bioinfo3d.cs.tau.ac.il/PatchDock/). The results refinement and energy calculation was performed as per the algorithm used in the Firedock server. In server, the final ranking was performed to identify the near-native refined solution. The ranking was based on various binding energy function that includes a variety of energy terms: van der Waals interactions, partial electrostatics, desolvation energy (atomic contact energy, ACE), hydrogen and disulfide bonds, p-stacking and aliphatic interactions and rotamer’s probabilities[24]. The result having minimum global energy was taken into consideration. Out of all 10 drugs predicted, Atazanavir (DrugBank ID – DB01072) (Figure 3a) has shown the minimum global energy. To acess the ligplots and detailed protein interactions, the solution were further analysed with PDBsum (http://www.ebi.ac.uk/thornton-srv/databases/cgi-bin/pdbsum/GetPage.pl?pdbcode=index.html) Figure 3b shows the ligplot analysis having conventional H-bonding between ligand and protein and 158 various non-bonded contacts and interctions. One H-bond was observed between Lys (12) A of the protein & O22 of the ligand. Similarly, the second one was between Lys(97)B & O7. These H-bonds favoured a strong bonding betwwen the two moieties. The bond lengths of the above two H-bonds are 3.07Å and 2.18Å respectively.

**Figure 3:**
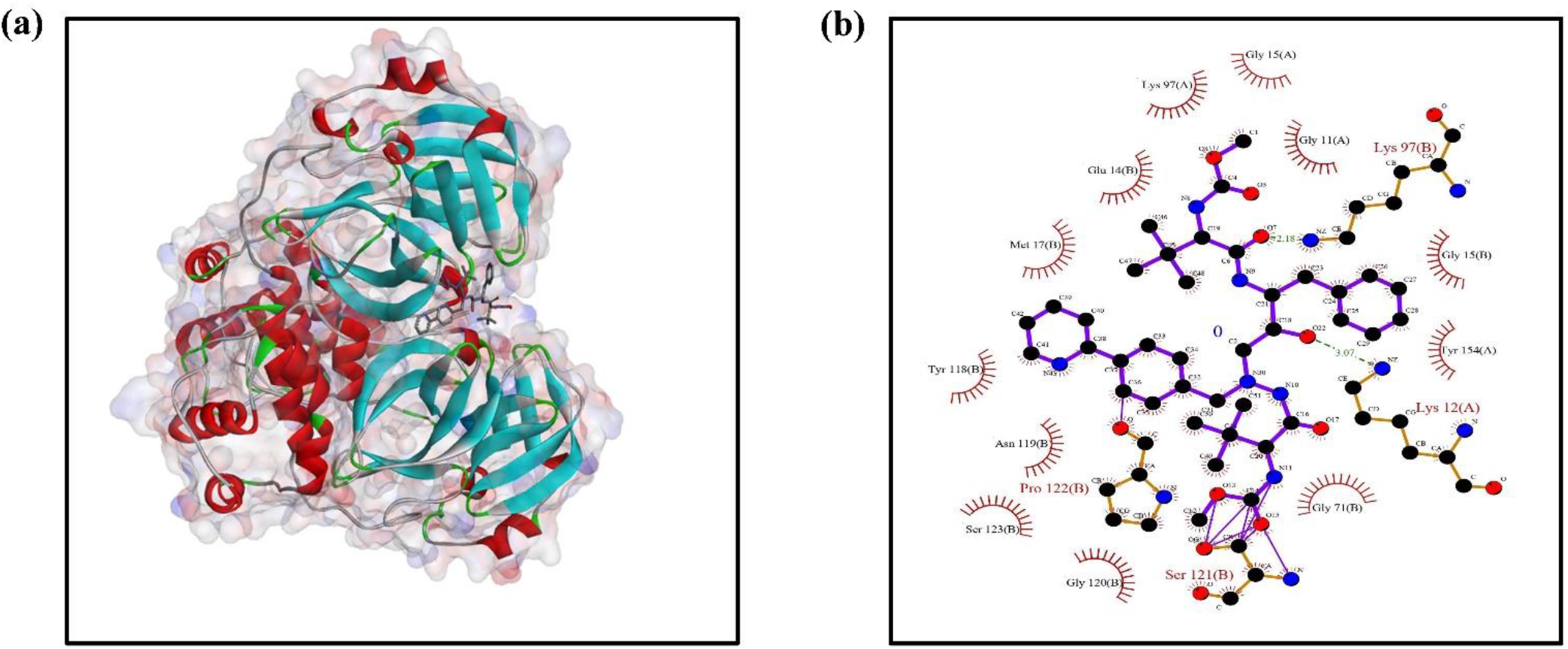
(a) Solution model showing Protein-ligand docking, (b) ligplot analysis showing possible bonds between protein and ligand.

## Discussion

In our study, we have taken those drugs for docking purpose which show protease inhibition activity. The docking of SARS protein with the approved drug Atazanavir (DrugBank ID – DB01072), has a global energy of around -59.90 Kcal/mol which confirms that it had numerous steric clashes with the adjoining strands and thus highlight its potential to inhibit novel SARS coronavirus. Atazanavir is a small molecule antiretroviral drug (Figure 2b), usually sold under the brand names Invirase and Fortovase, used together with other medications to treat or prevent HIV/AIDS. It is an HIV protease inhibitor which acts as an analog of an HIV protease cleavage site. It is a highly specific inhibitor of HIV-1 and HIV-2 proteases.

In addition to the docking of the above drug, we have also docked several other approved drugs available in the Drug Bank, which are predicted by our ML model with a confidence level of above 95% and also shows the activity of protease inhibition [Supplementary Information Figure S1-S9]. With reference to that, we have found that the other drugs predicted by the model with the inclusion of all the parameters taken under consideration can also be quite effective.

The rapid identification of active therapeutic agents against SARS-CoV-2 is a major challenge. Analyzing the available knowledge on their safety profiles, and in some cases, efficacy against other coronaviruses and repurposing existing antiviral drugs is a potentially crucial short-term strategy to tackle COVID-19.

Under the current scenario, it takes more than 15 years to bring a drug from the investigational stages to market availability. It is because of the trial and error process or the so-called Edisonian Approach, where one keeps on analyzing several compounds to find the best possible one. These days with the inclusion of digital medicine, this time span has been reduced to a great extent and people are able to approach in a rational manner for the drug discovery process. Here, we have targeted for the repurpssed drugs towards the development of effective treatment of COVID-19 to speed up clinical trial. We have found that ML model created on the basis of the Naive Bayes algorithm is the most effective one with the accuracy of almost equal to 73%. The drugs predicted by this model is further verified by the docking process. We speculate that our predicted drugs show immense potential for treatment of the COVID-19.

Considering the ongoing efforts to prevent the spread of COVID-19 all over the world, we are optimistic that the outbreak may subside in a few months like SARS and MERS. However, the outbreak has stressed the urgent need for renewed efforts towards the development of braod-spectrum therapeutic agents to combat coronaviruses which are repeatedly found to be a realistic threat of this century till now. Our this findinging will provide a base for further enhanced drug discovery programs.

## Data Availability

The data is available on request.

## Author Contribution

SM and PN have executed the machine learning part. MC and ND has executed the docking part. DK and PR have helped in writing and editing manuscript and medical inputs. SS has formulated the problem, instructed for execution method, mostly written and edited the manuscript.

## ACKNOWLEDGEMENTS

SS acknowledge Young Faculty Research Grant from Ministry of Information Science and Technology, Government of India.

## References

1. Perlman S. Another decade, another coronavirus. New England Journal of Medicine. 2020. doi:10.1056/NEJMe2001126

2. She J, Jiang J, Ye L, Hu L, Bai C, Song Y. 2019 novel coronavirus of pneumonia in Wuhan, China: emerging attack and management strategies. Clin Transl Med. 2020. doi:10.1186/s40169-020-00271-z

3. Graham Carlos W, Dela Cruz CS, Cao B, Pasnick S, Jamil S. Novel Wuhan (2019-NCoV) coronavirus. American Journal of Respiratory and Critical Care Medicine. 2020. doi:10.1164/rccm.2014P7

4. Gorbalenya AE, Baker SC, Baric RS, de Groot RJ, Drosten C, Gulyaeva AA, et al. The species Severe acute respiratory syndrome-related coronavirus: classifying 2019-nCoV and naming it SARS-CoV-2. Nat Microbiol. 2020. doi:10.1038/s41564-020-0695-z

5. Cui J, Li F, Shi ZL. Origin and evolution of pathogenic coronaviruses. Nature Reviews Microbiology. 2019. doi:10.1038/s41579-018-0118-9

6. Huang C, Wang Y, Li X, Ren L, Zhao J, Hu Y, et al. Clinical features of patients infected with 2019 novel coronavirus in Wuhan, China. Lancet. 2020. doi:10.1016/S0140-6736(20)30183-5

7. Zhou P, Yang X-L, Wang X-G, Hu B, Zhang L, Zhang W, et al. Discovery of a novel coronavirus associated with the recent pneumonia outbreak in humans and its potential bat origin. Nature. 2020. doi:10.1101/2020.01.22.914952

8. Paules CI, Marston HD, Fauci AS. Coronavirus Infections-More Than Just the Common Cold. JAMA - Journal of the American Medical Association. 2020. doi:10.1001/jama.2020.0757

9. Liu GS, Li H, Zhao SC, Lu RJ, Niu PH, Tan WJ. Viral and Bacterial Etiology of Acute Febrile Respiratory Syndrome among Patients in Qinghai, China. Biomed Environ Sci. 2019. doi:10.3967/bes2019.058

10. Dong Y, Mo X, Hu Y, Qi X, Jiang F, Jiang Z, et al. Epidemiological Characteristics of 2143 Pediatric Patients With 2019 Coronavirus Disease in China. Pediatrics. 2020. doi:10.1542/peds.2020-0702

11. Lu R, Zhao X, Li J, Niu P, Yang B, Wu H, et al. Genomic characterisation and epidemiology of 2019 novel coronavirus: implications for virus origins and receptor binding. Lancet. 2020. doi:10.1016/S0140-6736(20)30251-8

12. Wrapp D, Wang N, Corbett KS, Goldsmith JA, Hsieh CL, Abiona O, et al. Cryo-EM structure of the 2019-nCoV spike in the prefusion conformation. Science (80-). 2020. doi:10.1126/science.aax0902

13. Armstrong GL, MacCannell DR, Taylor J, Carleton HA, Neuhaus EB, Bradbury RS, et al. Pathogen genomics in public health. N Engl J Med. 2019. doi:10.1056/NEJMsr1813907

14. Gorbalenya AE. Severe acute respiratory syndrome-related coronavirus – The species and its viruses, a statement of the Coronavirus Study Group. bioRxiv. 2020. doi:10.1101/2020.02.07.937862

15. Huang R, Xia J, Chen Y, Shan C, Wu C. A family cluster of SARS-CoV-2 infection involving 11 patients in Nanjing, China. Lancet Infect Dis. 2020. doi:10.1016/s1473-3099(20)30147-x

16. Zhu N, Zhang D, Wang W, Li X, Yang B, Song J, et al. A novel coronavirus from patients with pneumonia in China, 2019. N Engl J Med. 2020. doi:10.1056/NEJMoa2001017

17. Tang B, Bragazzi NL, Li Q, Tang S, Xiao Y, Wu J. An updated estimation of the risk of transmission of the novel coronavirus (2019-nCov). Infect Dis Model. 2020. doi:10.1016/j.idm.2020.02.001

18. Melville J, Burke E, Hirst J. Machine Learning in Virtual Screening. Comb Chem High Throughput Screen. 2009. doi:10.2174/138620709788167980

19. Lowe R, Glen RC, Mitchell JBO. Predicting phospholipidosis using machine learning. Mol Pharm. 2010. doi:10.1021/mp100103e

20. Cheng T, Li Q, Zhou Z, Wang Y, Bryant SH. Structure-based virtual screening for drug discovery: A problem-centric review. AAPS Journal. 2012. doi:10.1208/s12248-012-9322-0

21. Mitchell TM. Machine Learning. Annual Review Of Computer Science. 1997. doi:10.1145/242224.242229

22. Schierz AC. Virtual screening of bioassay data. J Cheminform. 2009. doi:10.1186/1758-2946-1-21

23. Wang Y, Xiao J, Suzek TO, Zhang J, Wang J, Zhou Z, et al. PubChem’s BioAssay database. Nucleic Acids Res. 2012. doi:10.1093/nar/gkr1132

24. Mashiach E, Schneidman-Duhovny D, Andrusier N, Nussinov R, Wolfson HJ. FireDock: a web server for fast interaction refinement in molecular docking. Nucleic Acids Res. 2008. doi:10.1093/nar/gkn186

25. Wishart DS, Knox C, Guo AC, Cheng D, Shrivastava S, Tzur D, et al. DrugBank: A knowledgebase for drugs, drug actions and drug targets. Nucleic Acids Res. 2008. doi:10.1093/nar/gkm958

26. Protein Data Bank. RCSB PDB: Homepage. Rcsb Pdb. 2019.

27. Hop P, Allgood B, Yu J. Geometric Deep Learning Autonomously Learns Chemical Features That Outperform Those Engineered by Domain Experts. Molecular Pharmaceutics. 2017. doi:10.1021/acs.molpharmaceut.7b01144.

28. Ng A. 1. Supervised learning. Mach Learn. 2012. doi:10.1111/j.1466-8238.2009.00506.x

29. Friedman N, Geiger D, Goldszmidt M, Provan G, Langley P, Smyth P. Bayesian Network Classifiers *. Mach Learn. 1997. doi:10.1023/A:1007465528199

30. Refaeilzadeh P, Tang L, Liu. H. “Cross-Validation.” Encyclopedia of database systems. 2009. doi:10.1007/978-0-387-39940-9_565

31. Browne MW. Cross-validation methods. J Math Psychol. 2000. doi:10.1006/jmps.1999.1279

32. Sun Y, Kamel MS, Wong AKC, Wang Y. Cost-sensitive boosting for classification of imbalanced data. Pattern Recognit. 2007. doi:10.1016/j.patcog.2007.04.009

33. Thai-Nghe N, Gantner Z, Schmidt-Thieme L. Cost-sensitive learning methods for imbalanced data. Proceedings of the International Joint Conference on Neural Networks. 2010. doi:10.1109/IJCNN.2010.5596486

34. Wang J, Zhao P, Hoi SCH. Cost-Sensitive Online Classification. IEEE Trans Knowl Data Eng. 2014. doi:10.1109/TKDE.2013.157

35. Sen P, Getoor L. Cost-sensitive learning with conditional Markov networks. Data Min Knowl Discov. 2008. doi:10.1007/s10618-008-0090-5

36. Masnadi-Shirazi H, Vasconcelos N. Cost-sensitive boosting. IEEE Trans Pattern Anal Mach Intell. 2011. doi:10.1109/TPAMI.2010.71

